# Prioritising Follow-Up for People with Suspected Epilepsy Using a Digital EEG Biomarker

**DOI:** 10.1101/2025.06.30.25330550

**Authors:** Rosie Charles, Emanuela De Falco, Elizabeth Galizia, David Martin-Lopez, Kay Meiklejohn, David Allen, Lydia E. Staniaszek, Chris Price, Sophie Georgiou, Manny Bagary, Sakh Khalsa, Charlotte Lawthom, Rohit Shankar, John R. Terry, Wessel Woldman

**Author notes:** These authors contributed equally to this work as first authors (John R. Terry, Wessel Woldman). These authors contributed equally to this work as last authors Corresponding Author: Rosie Charles.

## Abstract

Lengthy waits for follow-up testing are common for people with suspected epilepsy. This delays diagnosis, prolongs uncertainty and increases seizure risk. Initial EEGs are frequently inconclusive, yet follow-ups are often dictated by referral date, and there is no established method for risk-based prioritisation. Here, we tested whether an established digital EEG biomarker could help prioritise those most likely to have epilepsy for expedited follow-up EEG testing. We analysed 196 normal non-contributory (non-diagnostic) initial EEGs collected from six National Health Service (NHS) sites in England. From these recordings, we extracted eight previously validated computational features that quantify the likelihood that the EEG was recorded from someone with active epilepsy. We then used this information to reorder follow-up lists and compared outcomes against standard referral-based scheduling.

We found that ordering for follow-up testing based upon the digital biomarker consistently prioritised people subsequently diagnosed with epilepsy; for a waitlist of 40 patients, the median number of follow-up EEGs needed to see 50% of true epilepsy patients was decreased by 6 (95% CI 4-7). The EEG diagnostic yield for epilepsy of follow-ups was increased relative to orderings based on time of referral (median increase in yield for epilepsy at 50% follow-up EEGs was 5%; 95 CI 4.9%-10%). Our study indicates that a routine EEG may furnish an objective risk metric that could accelerate second-line investigations and so reduce diagnostic delay whilst improving resource allocation in clinical practice.

## 1. Introduction

An audit of UK and Ireland seizure referral pathways [1] found that though routine EEGs are performed in over 50% of people with a suspected first seizure, the diagnostic yield of these EEGs is limited. A meta-analysis of 15 studies reporting the results of routine EEG in participants presenting with a first unprovoked seizure found the median sensitivity across studies for these first routine EEGs to be just 17.3% [2]. This presents a significant challenge where many first routine EEGs following a first seizure are inconclusive, and additional EEGs are required. From there, whilst individual hospital protocols vary, patients are largely allocated for further testing based on time of referral. However, it is important to note that in some clinical practices, discretionary judgement may be applied to prioritise patients who are found to be at highest risk. Consequent EEGs are usually ambulatory EEGs or sleep-deprived EEGs, with second routine EEGs comprising a small minority [3]. 17-34% of patients receiving a second EEG are eventually diagnosed with epilepsy following ambulatory EEG [4], compared to 29% for sleep-deprived EEG [5].

However, limited resources often result in long waits for further testing creating significant diagnostic delays and substantial risks. For example, an Australian review of seventeen studies reported that 13-16% of patients experienced diagnostic delays of 1 year or more [6]. Another retrospective study on newly diagnosed focal epilepsy patients at a Finnish epilepsy centre reported a median diagnostic delay of 12 months, extending to several years in some cases, with 77% experiencing more than two additional seizures before treatment began [7]. During diagnostic delay, many patients with experience a second seizure; a review of 58 studies with a total of 12,160 participants revealed that a quarter of adults who experience a first unprovoked seizure will experience another within six months, rising to 35% by twelve months [8]. Furthermore, the consequences of diagnostic delay extend beyond immediate seizure risk, affecting driving privileges, employment security, psychological wellbeing, and potentially compromising long-term prognosis through delayed access to appropriate treatment.

Recently, computational approaches have shown promise in enhancing diagnostic yield of clinical investigations. An Australia-based study found that scheduling video-EEG based on seizure-diary analysis significantly improved diagnostic yield from 50% to 62% (*p* < 0.001, >5000 video-EEGs), with confirmed seizures observed in 18% of high-risk cases versus 11% in standard scheduling [9].

Whilst seizure diaries offer valuable insights, their subjective nature is well documented [10]. More objective approaches have emerged through computational analysis of EEG data itself [11–16]. In previous work, a set of eight spectral-, network-and computer model-based features, otherwise hidden to standard visual EEG analysis, were developed [11]. At Neuronostics, we combined these features into a single computational biomarker of epilepsy: BioEP. BioEP achieved a sensitivity of over 60% for detecting epilepsy from non-contributory EEGs. Clinically non-contributory EEGs lack epileptiform activity, such as interictal epileptiform discharges or seizures, that would support a diagnosis of epilepsy. Generally speaking, they do not contain any spikes, sharp-waves or other patterns that hold diagnostic value for epilepsy or any of its differential conditions [17]. Non-contributory EEGs may further be classified as normal or abnormal [18–20]. An abnormal non-contributory EEG contains visually abnormal features non-specific to epilepsy, such as generalised delta slowing [18]. BioEP’s ’similarity score’ compares the eight features calculated from an individual’s non-contributory routine EEG with a database of people with known final diagnoses. Higher similarity scores indicate greater likelihood that the patient has epilepsy. In this study, we investigated a practical application for this biomarker: prioritisation of patients for follow-up EEG testing based on seizure risk as guided by BioEP’s ‘similarity score’.

Using 196 non-contributory first routine EEGs collected across six first seizure clinics, we evaluate whether score-guided prioritisation improves the diagnostic yield of epilepsy of follow-up EEGs compared to time-of-referral allocation. Our findings suggest that this approach ensures people ultimately diagnosed with epilepsy are seen sooner. This pragmatic integration of digital innovation with clinical practice potentially enhances diagnostic efficiency, reduces risks of uncontrolled seizures, and lowers costs associated with delayed diagnosis.

## 2. Methods

### 2.1 Dataset

196 subjects with normal non-contributory first routine EEGs were selected from a total population of 281 individuals with both normal and abnormal non-contributory EEGs. This dataset was previously collated in a retrospective, multi-site study (see Tait et al. (2024) [11], and https://www.clinicaltrials.gov/study/NCT05384782). Exclusion criteria for the set included subjects under 18 years of age, clinically contributory EEGs, EEGs containing non-epileptiform abnormalities, EEGs in non-compatible data formats, and subjects with missing metadata. Inclusion criteria required a first awake EEG, and that subjects have a final diagnosis (epilepsy or otherwise) which had remained stable for at least one year. For all patients, the diagnosis was not made at the time of their first non-contributory EEG but at some point thereafter; in other words, at the point of their first available EEG, there was a very high degree of diagnostic uncertainty [2,21] as to whether the subject actually had epilepsy, or a single provoked seizure, or a differential condition such as functional/dissociative seizures (FDS) [22].

A breakdown of the characteristics of this cohort is reported in Table 1. All EEGs were from unique subjects who had reported experiencing at least one suspected seizure and who were being investigated for possible epilepsy. Sixty-six subjects were eventually diagnosed with epilepsy (‘Ep’, includes subjects with focal and generalised seizures) after follow-up testing, and 130 subjects eventually had a diagnosis of epilepsy ruled out following further clinical assessment, in line with standard practice (‘NEp’, includes cases of syncope and FDS). This distribution is consistent with the typical prevalence of epilepsy in individuals undergoing a second EEG [4]. Abnormal non-contributory EEGs, such as those including subtle abnormalities such as focal slowing, asymmetries and background disorganization were not included in the selected dataset from either Ep or NEp subjects. Formal sample-size calculations were not required due to the fact this is study considers the application of an established set of markers and model (e.g. as opposed to a biomarker development study). The narrow confidence intervals in our primary outcome measures indicate the number of subjects considered are appropriate.

**Table 1.** Summary of patients’ characteristics

| Characteristic | Category | Value |
| --- | --- | --- |
| Age; Mean (SD; range), years |  | 38.8 (14.8; 18–91) |
| Sex n (%) | Female | 105 (53.6) |
|  | Male | 90 (45.9) |
|  | Other | 1 (0.5) |
| ASM Treatment Status, n (%) | Yes | 74 (37.8) |
|  | No | 122 (62.2) |
| Epilepsy diagnosis, n (%)<br>Total: 66 (33.7) | Focal | 50 (75.7) |
|  | Generalised | 5 (7.6) |
|  | Unknown | 11 (16.7) |
| Non-Epilepsy diagnosis, n (%)<br>Total: 130 (66.3) | Functional (Dissociative) Seizures | 64 (49.2) |
|  | Syncope | 15 (11.5) |
|  | Movement Disorder | 6 (4.6) |
|  | Sleep Disorder | 3 (2.3) |
|  | Other/Unclassified | 42 (32.3) |

### 2.2 Analysis

All analyses were performed using custom Python scripts. Pre-processing of EEG was performed using MNE-Python (v1.6.1) [23]. Normality of the distributions was assessed using the Shapiro-Wilk test. For most distributions, the test indicated a significant deviation from normality (p < 0.05); therefore, all statistical group comparisons were performed using the Wilcoxon signed-rank test from the scipy.stats module [24].

### 2.3 EEG preprocessing and feature extraction

EEGs were recorded using the international 10–20 system. We included 19 channels (Fp1, Fp2, F7, F3, Fz, F4, F8, C3, Cz, C4, P3, Pz, P4, T3, T4, T5, T6, O1, O2). A maximum of 20 minutes of EEG recording was included for each subject (range: 15.8-20 minutes, mean ± SD: 19.9 ± 0.4 minutes). EEGs were segmented into 20-second epochs, and epochs containing artefacts were excluded. Eight features were then extracted, as described in previ-ous work [11], comprising two spectral, four network-based, and two model-based features (detailed in Table S1). Briefly, spectral features capture alpha-band characteristics, network features measure overall properties of the communication between brain regions, while model-based features reflect dynamical properties of the EEG derived from a mathematical model parametrised using the EEG data. Full feature definitions, computation details, and value ranges are provided in the Supplementary Material.

### 2.4 Confounder Model

Some patient characteristics and medication-related factors can systematically influence EEG features and, therefore, must be accounted for to ensure that our methodology is robust to potential confounding effects. Following previous findings [11], we accounted for confounding effects of sex and anti-seizure medication (ASM) status, both of which have been shown to significantly influence the eight EEG features used in our methodology. For each extracted feature, confounder correction was applied using β coefficients derived from generalised linear models (GLMs) fitted to the training sets:

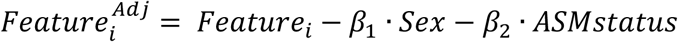

Here *Featurei* represents one of the eight features, *Sex* is a categorical variable (0: female; 1: male; 2: other) and *ASMstatus* is a binary variable (0: not taking ASMs; 1: taking ASMs at time of EEG). Adjusted features were then used in all the subsequent analysis steps.

### 2.5 Prioritisation Based on Classifier Confidence

A random undersampling boosting (RUB) model with four principal components has previously been identified as the best-performing classifier of epilepsy [11]. The classifier takes the eight adjusted features as its inputs. For this study, we used a fixed partition: 106 EEGs (Ep = 36, NEp = 70) for training and 90 EEGs (Ep = 30, NEp = 60) for testing, with matching Ep:NEp ratios (Figure 1). The fixed partitioning approach ensures that the EEG of each individual in the testing set receives a single similarity score produced by a single model.

**Figure 1.**
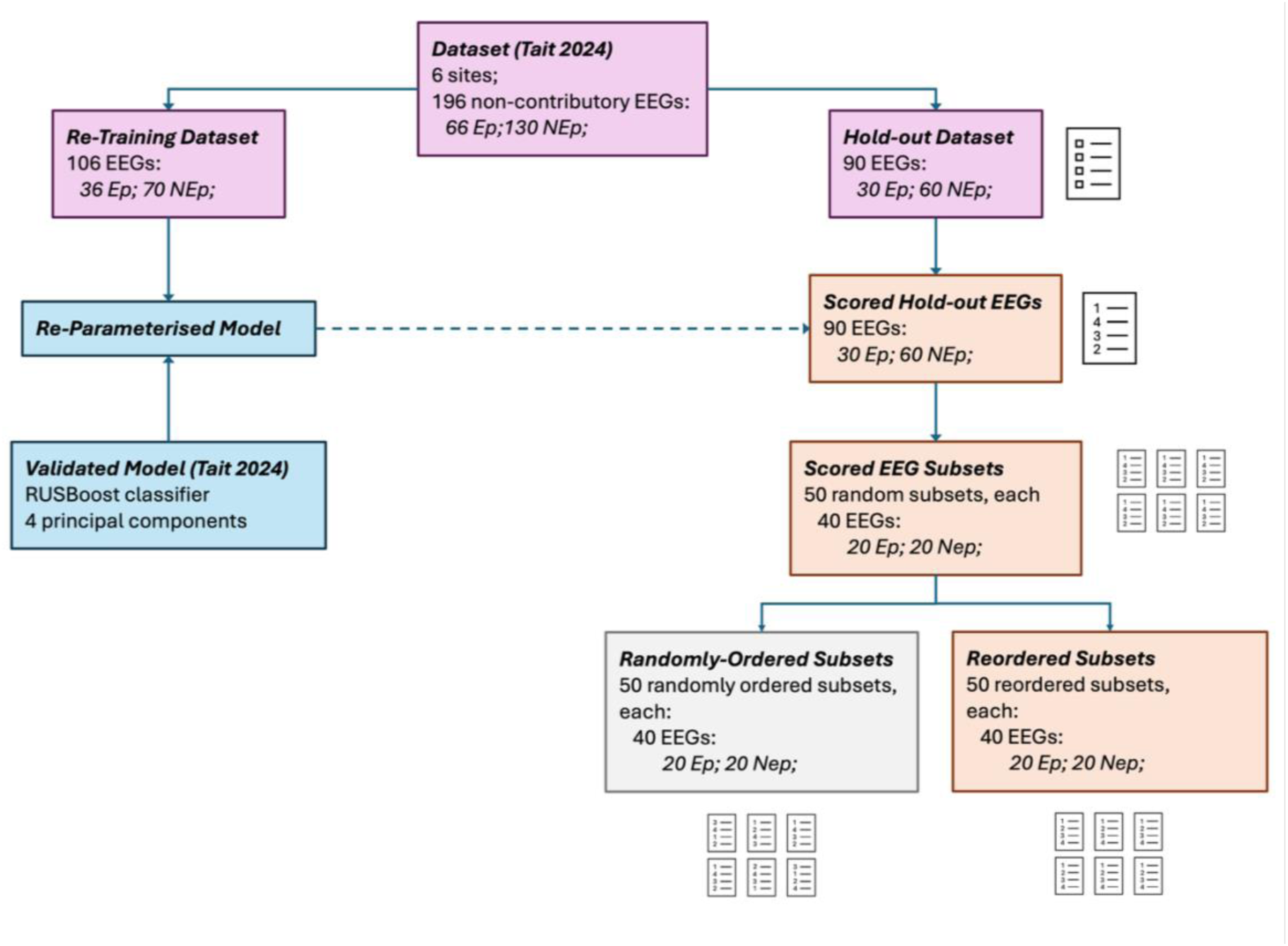
Dataset profile and prioritisation pipeline. Flow diagram summarising the partitioning of the data, retraining of the previously validated RUB model and scoring and reordering of a hold-out set using this model.

The trained RUB model produced a score between 0 and 1 for each individual in the testing set. This score represents the confidence of the classifier that a given EEG originated from an individual with active epilepsy, and as such is termed the “similarity score”.

To simulate clinical prioritisation, we drew 50 random subsets of 40 individuals from the 90 EEG test-set, with a 50:50 Ep:NEp ratio for each subset. This balanced ratio was chosen as a baseline scenario against which to evaluate our method. In each subset individuals were prioritised based on their classifier scores, from highest to lowest, to obtain an ordered list for follow-up appointments. For comparison, we also generated a randomised order for each subset akin to a standard referral-based scheduling approach.

We repeated this process for subsamples in which the prevalence of epilepsy varied from 10-80% to assess generalisability across different clinical settings.

### 2.6 Impact on Clinical metrics

To assess the potential impact of biomarker-based prioritisation on clinical metrics, we envisioned a test scenario where our patients, which all had an inconclusive first EEG and persistent diagnostic uncertainty, were scheduled for a follow-up prolonged EEG (either ambulatory or sleep-deprived EEG), which is one of the standard clinical pathways in cases of uncertain diagnosis and negative routine EEG. We then evaluated the impact of prioritisation of those follow-up visits by comparing biomarker-ordered and random-ordered lists on several metrics. First, we calculated the number of follow-up EEGs required to be completed to have seen 15%, 50%, and 80% of people with epilepsy. We then assessed cumulative percentage of Ep and NEp individuals seen, and the cumulative ‘EEG diagnostic yield for epilepsy’ as a function of follow-up EEGs performed on each ordered list. The ‘EEG diagnostic yield for epilepsy’ here represents the proportion of follow-up EEGs which are expected to contain epileptiform activity and was used to estimate how biomarker-based prioritisation might increase the likelihood that patients with epilepsy reach a diagnostically useful EEG earlier in their pathway.

To estimate diagnostic yield of the follow-up EEG for epilepsy we assumed an average sensitivity of 54% for second-line EEG investigations. This was estimated based on the sensitivity of the most common second-EEG types performed following a first routine EEG: ambulatory EEGs (sensitivity: 63%) and sleep-deprived EEGs (sensitivity: 45%) [3,25].

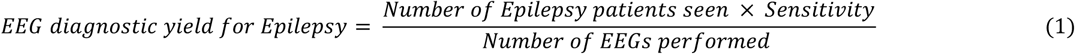

which represents the proportion of follow-up EEGs that contains epileptiform activity and therefore supports a diagnosis of epilepsy.

## 3. Results

We first assessed how effectively the biomarker prioritised people with epilepsy by estimating the number of EEGs required to have seen 15%, 50% and 80% of Ep patients across the 50 ordered subsamples, each composed of 40 patients (Figure 2). These values were compared to those obtained from the randomly ordered lists using the Wilcoxon signed-rank test. Biomarker-based ordering significantly reduced the number of EEGs needed to reach each threshold, indicating improved prioritisation of individuals with epilepsy. For example, the median improvement in number of follow-up EEGs needed to see 50% of Ep patients was 6 (95% CI: 4-7, effect size: 0.83). Full statistical results are reported in Supplementary materials section (Table S2).

**Figure 2.**
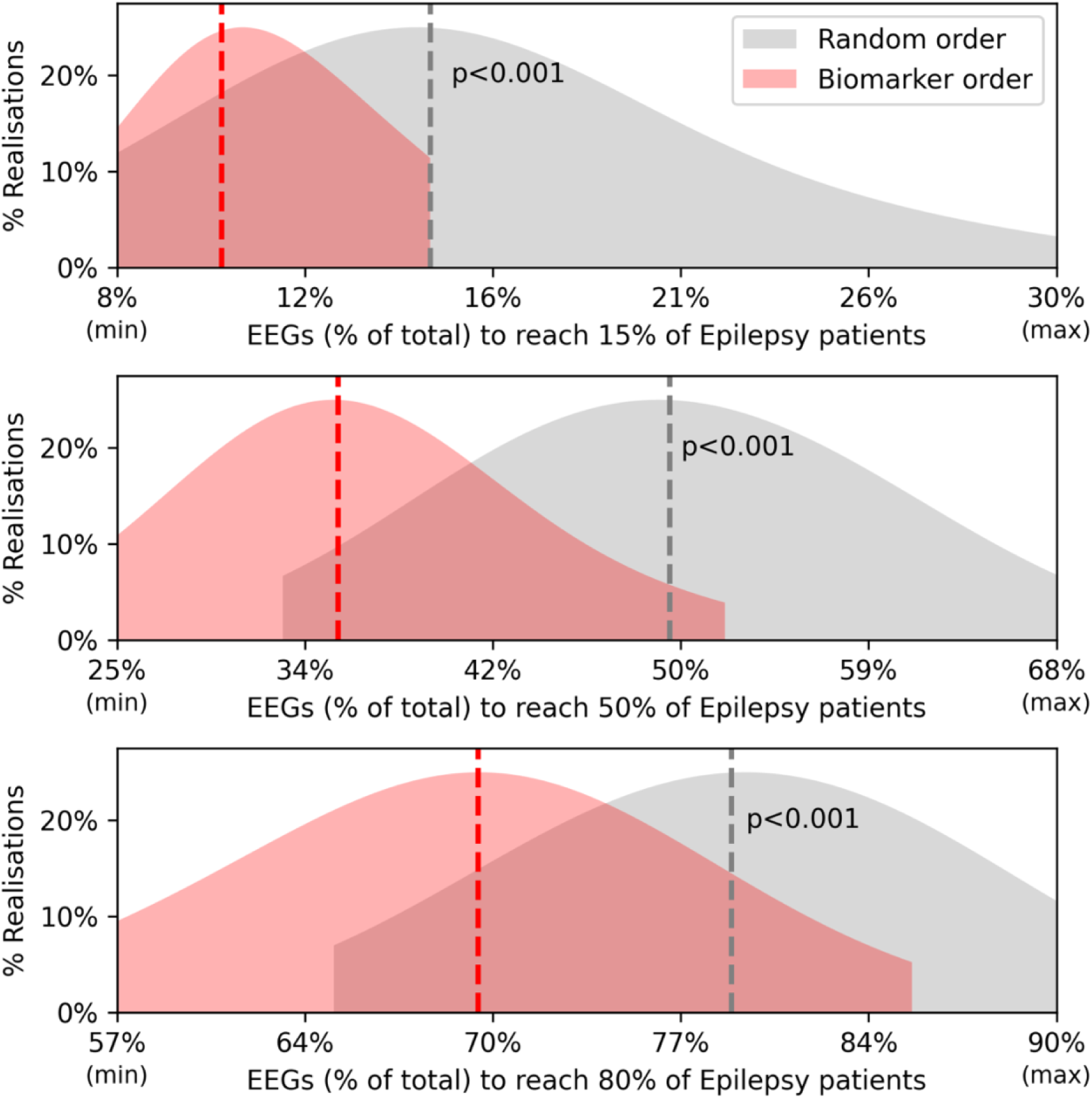
Numbers of EEGs needed to reach specific percentages of people with epilepsy receiving a follow-up EEG. Red curves show the distributions of EEGs required to reach 15%, 50% and 80% of Ep individuals across the 50 realisations. Grey curves represent the values obtained from the random distribution. Dotted vertical lines represent median of each distribution. p-values reflect statistical comparisons. The extremes of the x-axis correspond to the observed minimum and maximum values.

To evaluate practical benefit of our re-ordering, we plotted the cumulative percentages of individuals seen (Ep and NEp) as a function of the percentage of EEGs performed (Figure 3.A1). This shows that our method successfully prioritises patients with epilepsy, with a consistently higher proportion of people with epilepsy appearing in the upper portion of a re-ordered list. In addition, we estimated the EEG diagnostic yield for epilepsy following the biomarker-based reordering against random ordering (Figure 3.A2). Biomarker-based prioritisation consistently yielded improvements in diagnostic efficiency compared to random scheduling, above what would be expected by chance.

**Figure 3.**
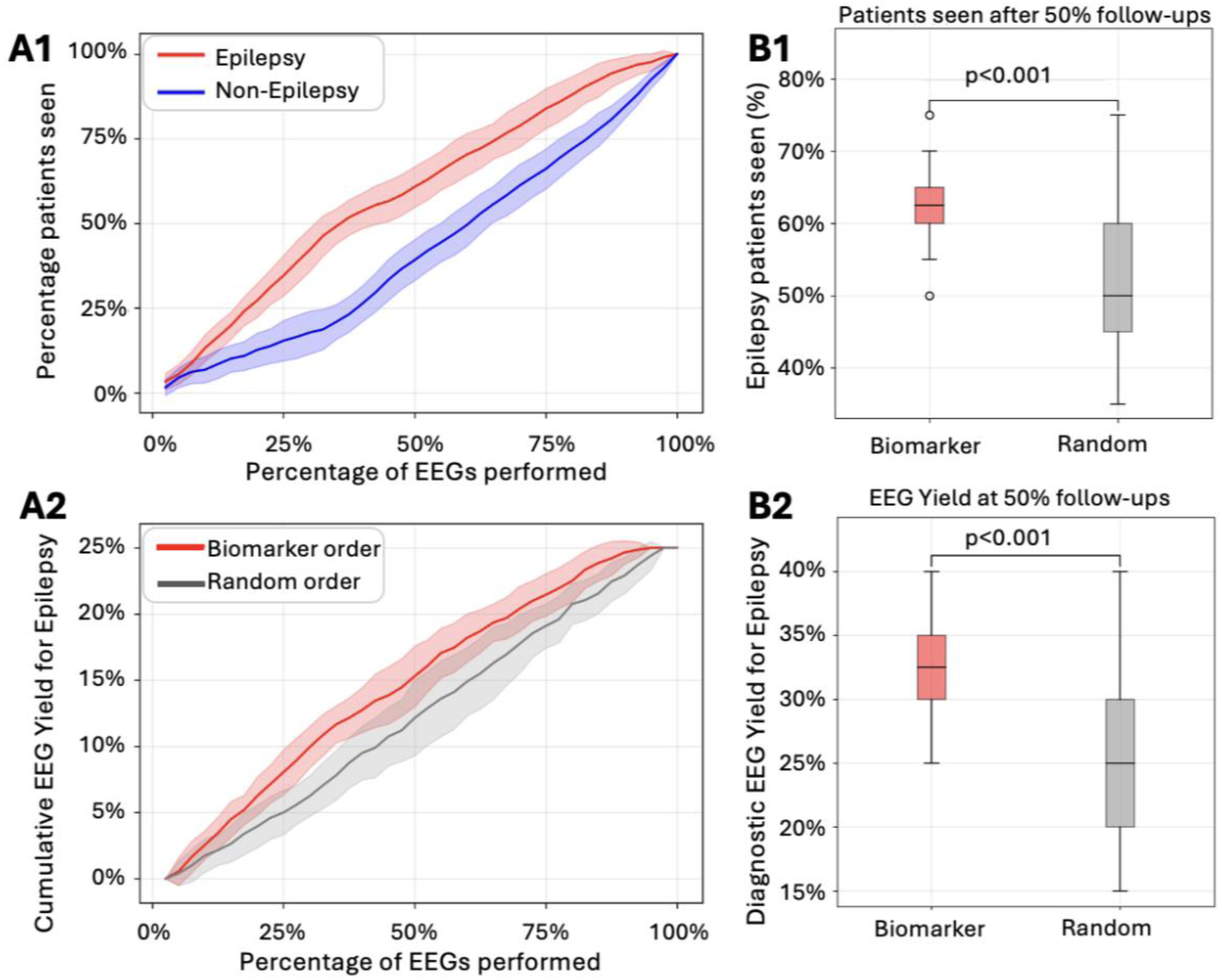
A1) Percentage of Ep (red) and NEp (blue) individuals seen versus percentage of EEGs performed. **A2)** EEG Diagnostic yield for epilepsy over time versus percentage of EEGs using the classifier ordering (red) and random scheduling (grey). Solid lines indicate mean value across the 50 realisations; shaded areas indicate standard deviations. **B1)** Number of people with epilepsy seen after 50% of the EEGs (biomarker-based (red) random scheduling (grey). **B2)** Corresponding EEG diagnostic yield for epilepsy at 50% follow-up EEGs. Boxplots indicate median, and interquartile range. p-values reflect statistical comparisons.

We next examined the midpoint impact: the number of people with epilepsy seen within the first 50% of the follow up visits (Figure 3.B1), and the corresponding EEG diagnostic yield for epilepsy (Figure 3.B2). In both cases, biomarker-based ordering significantly outperformed random scheduling (Wilcoxon signed-rank test). The proportion of Ep patients seen after 50% follow-up visits increased by 10% (median value, 95% CI 7-15%, effect size 0.76). The corresponding increase in yield for epilepsy was 5%, 95 CI 4.9%-10%, effect size 0.72). These highlight the potential of biomarker-based scheduling for earlier diagnosis and treatment.

To explore robustness across different real-world scenarios, we repeated the above subsampling and reordering process using samples with varying epilepsy prevalence (10% to 80%) (Figure S1). The benefits of biomarker-based ordering were most pronounced when epilepsy prevalence was low to moderate (10–50%), which is closer to the typical prevalence observed in clinics. As expected, the marginal gain diminished as prevalence increased, given that a higher proportion of epilepsy cases results in a higher likelihood of capture by chance under random ordering.

## 4. Discussion

### 4.1 Significance

Our findings demonstrate that a digital EEG biomarker applied to non-contributory routine EEGs can be used to improve the prioritisation of follow-up testing for people with suspected epilepsy. By reordering individuals using a biomarker-derived similarity score, individuals with epilepsy are seen earlier, resulting in a higher EEG diagnostic yield for epilepsy than seeing people based on time of referral. This has the potential to expedite diagnosis, reduce seizure burden, and improve resource allocation within stretched healthcare systems [7].

It is important to emphasise that this study represents a case study of a biomarker application rather than a biomarker development study. The biomarker and classifier were developed and validated as reported in Tait et al. (2024) [11], which was underpinned by a statistical framework including formal sample-size calculations for candidate features. Cross-validation of this classifier was performed in the original development study and is therefore not required for this application study (which uses a pre-validated tool). Instead, the final model from Tait et al. (2024) was re-parameterised on the relevant data for this application. This study represents an important intermediate step between the development of a biomarker and its potential clinical implementation. In future studies, other clinical metrics could be directly considered, including broader patient and system level outcomes that cannot be assessed retrospectively. For example, biomarker-guided prioritisation may reduce time to diagnosis for people with epilepsy, thereby shortening the time to treatment initiation and the occurrence of uncontrolled seizures, and generally affecting the seizure outcome [7,26]. At a more systemic level, one could evaluate its impact on rates of secondary and emergency admissions and overall healthcare utilisation and costs. Broader patient quality of life indicators may also be relevant, such as the psychological burden and socio-economic consequences of diagnostic delays [27]. Assessing these outcomes prospectively would provide a more comprehensive evaluation of the ultimate clinical utility of the biomarker.

It must be noted that our study has inherent limitations that should be explored in carefully designed prospective studies. Whilst the classifier was trained on hundreds of EEGs, expanding the training data to reflect larger and more heterogeneous populations is necessary, in particular with respect to rarer types of epilepsy. We also assumed uniform diagnostic yields for follow-up EEG types; real-world data will be important to confirm the robustness of this assumption. Within our cohort, diagnostic uncertainty was present given the non-contributory nature of the EEG. However, we cannot be certain that every individual (both Ep and NEp) had a follow-up EEG. Furthermore, our randomized approach did not factor in how identified clinical risks might impact clinical decision making (e.g. severity / frequency of events, history of seizures during childhood), and it is important to note that clinical expertise should always take precedence. In future studies, hybrid models should be developed that integrate clinical risk factors, existing survey-triage tools [28], and EEG based biomarkers. Finally, the model was calibrated such that classification of those most likely to have epilepsy were prioritised. We should emphasise that individuals with non-epileptic causes of suspected seizures are of equal importance clinically, and the same approach could be utilised to stimulate expedited psychiatric or cardiac testing for the subjects with the lowest estimated likelihood of epilepsy.

### 4.2 Conclusion

In conclusion, integration of digital technologies into the diagnostic pathway can offer a pragmatic, data-guided approach to addressing the challenge of epilepsy diagnosis in resource limited settings. These findings support the development of prospective clinical studies to test the real-world implementation of EEG biomarker-guided prioritisation of follow-up EEGs in settings where resources are limited. Such studies should consider the clinical usability, specific patient outcomes, and cost-effectiveness of implementation of this use-case into the existing workflow. Prioritisation of those most in need, without excluding others from follow-up, should enhance and not replace clinical judgment.

## Funding Information

Funding was provided by Innovate UK, National Institute for Health and Care Research, Engineering and Physical Sciences Research Council, and Epilepsy Research UK.

## Ethics Statement

This study was approved by the HRA & HCRW (Integrated Research Application System (IRAS)24: 260729) and by the internal ethics committees of all collaborating clinical institutes.

## Data Availability

The EEG recordings and metadata are not publicly available due to restrictions by privacy laws. Postprocessed data supporting the findings of this study are available upon reasonable request from the corresponding author, and additional metadata may be made available upon reasonable request from the study sponsor (R.S.).

## Acknowledgements

W.W. was supported by Epilepsy Research UK (F2002), Innovate UK (103939), and the NIHR (AI01646). J.R.T. was supported by the EPSRC (EP/N014391/2 and EP/T027703/1), Innovate UK (103939), and the NIHR (AI01646). R.S. was supported by Innovate UK (103939) and the NIHR (AI01646). We thank Anke Verhaege and Amy Amin for comments on an earlier draft of the manuscript.

## Author Contributions (CRediT)

**Rosie Charles:** Conceptualization, Methodology, Validation, Formal Analysis, Writing – Original Draft. **Emanuela De Falco:** Conceptualization, Methodology, Validation, Formal Analysis, Writing – Original Draft. **Elizabeth Galizia:** Investigation, Data Curation. **David Martin-Lopez:** Investigation, Data Curation. **Kay Meiklejohn:** Investigation, Data Curation, Writing – Review & Editing. **David Allen:** Investigation, Data Curation. **Lydia E. Staniaszek:** Investigation, Data Curation, Writing – Review & Editing. **Chris Price:** Investigation, Data Curation. **Sophie Georgiou:** Investigation, Data Curation. **Manny Bagary:** Investigation, Data Curation. **Sakh Khalsa:** Investigation, Data Curation. **Charlotte Lawthom:** Investigation, Data Curation. **Rohit Shankar:** Writing – Review & Editing, Supervision. **John R. Terry:** Conceptualization, Supervision, Writing – Review & Editing**. Wessel Woldman:** Validation, Conceptualization, Supervision, Writing – Review & Editing.

## Declaration of Competing Interests

Rosie Charles, Emanuela De Falco and Kay Meiklejohn are employees of Neuronostics. Wessel Woldman and John R. Terry are cofounders, directors, and shareholders of Neuronostics. Rohit Shankar has received institutional research, travel support and/or honorarium for talks and expert advisory boards from LivaNova, UCB, Eisai, Veriton Pharma, Bial, Angelini, UnEEG and Jazz/GW pharma outside the submitted work. He holds or has held competitive grants from various national grant bodies including Innovate, Economic and Social Research Council (ESRC), Engineering and Physical Sciences Research Council (ESPRC), National Institute of Health Research (NIHR), NHS Small Business Research Initiative (SBRI) and other funding bodies including charities all outside this work. He has worked in a research capacity with Neuronostics as chief investigator for 3 projects including the project from which the data for this paper was derived.

None of the other authors have any conflict of interest to disclose. We confirm that we have read the Journal’s position on issues involved in ethical publication and affirm that this report is consistent with those guidelines.

## Notes

### Competing Interest Statement

R.C., E.D.F. and K.M. are employees of Neuronostics. W.W. and J.R.T. are cofounders, directors, and shareholders of Neuronostics. R.S. has received institutional research, travel support and/or honorarium for talks and expert advisory boards from LivaNova, UCB, Eisai, Veriton Pharma, Bial, Angelini, UnEEG and Jazz/GW pharma outside the submitted work. He holds or has held competitive grants from various national grant bodies including Innovate, Economic and Social Research Council (ESRC), Engineering and Physical Sciences Research Council (ESPRC), National Institute of Health Research (NIHR), NHS Small Business Research Initiative (SBRI) and other funding bodies including charities all outside this work. He has worked in a research capacity with Neuronostics as chief investigator for 3 projects including the project from which the data for this paper was derived.
None of the other authors have any conflict of interest to disclose. We confirm that we have read the Journal's position on issues involved in ethical publication and affirm that this report is consistent with those guidelines.

### Author Declarations

This study was approved by the HRA & HCRW (IRAS ID: 260729) and by the R&D of all collaborating clinical institutes. The study was exempt from REC review as research is limited to use of previously collected, non-identifiable information.

### Summary of Updates

Revised following comments from journal reviewers

