## Supplementary for "Prioritising Follow-Up for People with Suspected Epilepsy Using a Digital EEG Biomarker"

### Supplementary material

#### *Effect of prioritization in different scenarios with varying diagnostic ratios*

The average prevalence of epilepsy patients undergoing a second EEG is estimated to be around 35%[1], but large variations are expected depending on the site. Therefore, we repeated our subsampling and re-ordering procedure while varying the prevalence of true epilepsy patients in the samples from 10% to 80% of the sample. For this analysis, we used a sample size of 30 patients to ensure variability in patients across our samples, and 200 repetitions.

For varying composition of the samples, we measured the 'relative increase in EEG diagnostic yield for epilepsy' as the EEG diagnostic yield obtained following the biomarker-based re-ordering divided by the expected yield given the sample composition and a sensitivity for epilepsy of 54%. We found that our reordering approach performs optimally during the first 50% of follow-up tests at lower prevalence of epilepsy as when prevalence of epilepsy is low, the current "real-life" approach is much less likely to accidentally prioritise epilepsy patients than when prevalence is high.

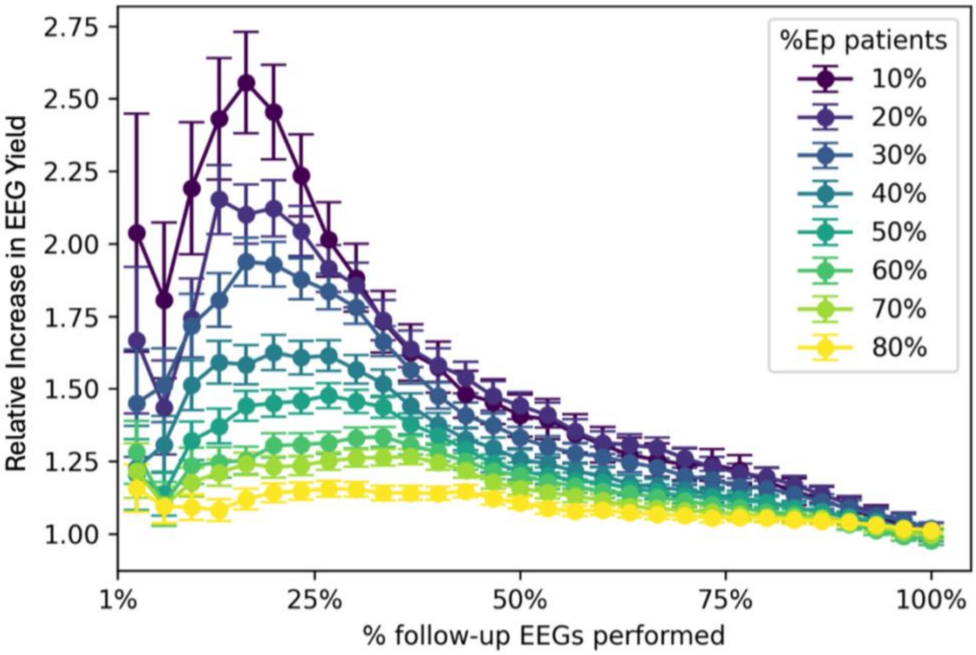


***Figure S1**. Effect of re-ordering on relative diagnostic yield in different scenarios. Relative increase in EEG yield as a function of the fraction of follow-up EEGs performed. For each scenario (with varying prevalence of Ep patients), the subsampling and re-ordering procedures were carried out, and the relative increase in yield was calculated at each step by dividing the measured yield by the expected yield given the sample composition.*

#### *EEG features*

The eight EEG features used to derive our biomarker were originally introduced and motivated across multiple prior studies[2–6] where they were shown to be relevant for distinguishing epilepsy from non-epilepsy. These features were subsequently consolidated and jointly validated into a biomarker in Tait et al. (2024)[7]. Together, they capture multiple aspects of EEG activity, including spectral characteristics, network connectivity, and model-based dynamical properties.

Spectral features comprised the peak alpha frequency, computed from occipital electrodes (O1 and O2) within the 8–13 Hz band, and the normalised alpha power, measured between frontal and temporal channels after bipolar montage was applied. Network features measured global properties of the communication between brain regions. To compute them, we first measured the connectivity for each pair of electrodes across all 19 channels (using the phase locking value in the 6-9Hz band) and then extracted four properties of this network: mean degree, degree variance, average weight clustering coefficient and characteristic path length. Model-based features were derived from a mathematical model of EEG parametrised using the EEG data and include: the local coupling, measuring the maximum local synchronisation, and the critical coupling measuring the theoretical threshold that can cause synchronisation across all electrodes. All features were corrected for potential confounders (see Methods) before being used in the classifier.

| Feature Name | Feature type | Description | Electrodes | Range |
|---|---|---|---|---|
| **Peak Alpha Frequency[2]** | Spectral | Frequency at which alpha power peaks | O1, O2 | [8,13] |
| **Alpha Power[3]** | Spectral | Relative power in the 10.5–13.5-Hz band | Frontal and temporal (bipolar montage) | [0,1] |
| **Mean degree[4,6]** | Network | Average number of connections per node | All 19 EEG channels | [0,18] |
| **Degree variance[4,6]** | Network | Variability of degree number across network | All 19 EEG channels | [0,5]* |
| **Average Weighted Clustering Coefficient[4,6]** | Network | Measures the tendency of the network to form clusters of connected nodes | All 19 EEG channels | [0,1] |
| **Characteristic Path Length[4,6]** | Network | Average shortest path between pairs of neds in the network | All 19 EEG channels | [1,9]* |
| **Local coupling[5,6]** | Model-based | Maximum local synchronisation from EEG-parametrised model | All 19 EEG channels | [0,0.3]* |
| **Critical coupling[5,6]** | Model-based | Threshold at which global synchronisation occurs in the EEG-parametrised model | All 19 EEG channels | [0,0.8]* |

***Table S1. EEG features underlying the Biomarker. The range indicates the theoretical range when this is defined and the typical observed range in EEG networks for the other cases (marked by asterisk). References next to the feature name refer to the relative discovery papers.***

#### *Table S2: Statistical results*

For all statistical comparisons reported, we used the Wilcoxon signed-rank test with a two-sided alternative hypothesis and sample sizes of N=50. A s a measure of effect size we used the absolute rank-biserial correlation (|*r*|).

| Comparison | Median difference | Confidence interval | T (signed rank sum) | p-value | Z-score | Effect size (\|*r*\|) |
|---|---|---|---|---|---|---|
| EEGs required to reach specific % of Ep individuals biomarker-based order vs random (Fig.2) | | | | | | |
| 15% of Ep patients | 2.0 | (1.0, 2.0) | 91.5 | 1.17E-06 | -5.27 | 0.75 |
| 50% of EP patients | 6.0 | (4.0, 7.0) | 27.5 | 5.62E-09 | -5.89 | 0.83 |
| 80% of EP patients | 3.5 | (3.0, 6.0) | 68 | 1.43E-07 | -5.50 | 0.78 |
| Epilepsy patients seen after 50% of follow-ups (Fig. 3-B1) | 0.1 | (0.07, 0.15) | 83.5 | 5.47E-07 | -5.35 | 0.76 |
| EEG Diagnostic yield for epilepsy after 50% of follow-ups (Fig. 3-B2) | 0.05 | (0.049, 0.1) | 109.5 | 1.71E-06 | -5.10 | 0.72 |

***Table S2. Statistical results***
